# Examination of the coverage of functional assessments in the OMOP common data model

**DOI:** 10.1101/2024.05.03.24306841

**Authors:** Margaret A. French, Paul Hartman, Heather A. Hayes, Leah Ling, John Magel, Anne Thackeray

## Abstract

**Background:** High-value care aims to enhance meaningful patient outcomes while reducing costs. Curating data across healthcare systems with common data models (CDMs) would help these systems move towards high-value healthcare. However, meaningful patient outcomes, such as function, must be represented in commonly used CDMs, such as Observational Medical Outcomes Partnership Model (OMOP). Yet the extent that functional assessments are included in the OMOP CDM is unclear.

**Objective:** Examine the extent that functional assessments used in neurologic and orthopaedic conditions are included in the OMOP CDM.

**Methods:** After identifying functional assessments from clinical practice guideline, two reviewer teams independently mapped the neurologic and orthopaedic assessments into the OMOP CDM. After this mapping, we measured agreement with the reviewer team with the number of assessments mapped by both reviewers, one reviewer but not the other, or neither reviewer. The reviewer teams then reconciled disagreements, after which we again examined agreement and the average number of concept ID numbers per assessment.

**Results:** Of the 81 neurologic assessments, 48.1% were initially mapped by both reviewers, 9.9% were mapped by one reviewer but not the other, and 42% were unmapped. After reconciliation, 46.9% of neurologic assessments were mapped by both reviewers and 53.1% were unmapped. Of the 79 orthopaedic assessments, 46.8% were initially mapped by both reviewers, 12.7% were mapped by one reviewer but not the other, and 48.1% were unmapped. After reconciliation, 48.1% of orthopaedic assessments were mapped by both reviewers and 51.9% were unmapped. Most assessments that were mapped had more than one concept ID number (neurologic assessments: 2.2±1.3; orthopaedic assessments: 4.3±4.4).

**Conclusions:** The OMOP CDM includes a portion of functional assessments recommended for use in neurologic and orthopaedic conditions. Many assessments did not have any term in the OMOP CDM. Thus, expanding the OMOP CDM to include recommended functional assessments and creating guidelines for mapping functional assessments would improve our ability to harmonize these data across healthcare systems.

## 1. BACKGROUND AND SIGNIFICANCE

The United States spends twice as much annually per person on healthcare compared to other high-income countries, yet we obtain worse health outcomes (e.g., shorter life expectancy, lower quality of life).[1, 2] This necessitates a shift towards high-value healthcare, where meaningful patient outcomes are improved while lowering the costs associated with these outcomes (i.e., better outcomes per dollar spent).[3–5] Large real-world data about interventions, outcomes, and costs are needed to move towards high-value care because they allow us to identify what interventions result in the best outcomes at the lowest cost. The outcomes needed for value-based care initiatives must be meaningful to patients.[3–6] While there are a number of important outcomes, function is particularly important because it is salient across all health diagnoses.[3–6] Thus, large-scale real-world databases that include assessments of function, which are routinely collected by rehabilitation professionals, are needed to improve the value of healthcare.

The widespread adoption of electronic medical records (EMRs) has allowed for the generation of these types of real-world databases. These real-world databases are even more useful for value-based care initiatives when data is aggregated across healthcare systems. Unfortunately, the same metric, such as a functional assessment, is frequently referred to differently across healthcare systems (Table 1, *Healthcare System 1 and 2 columns*), presenting a barrier to aggregating data across systems. To overcome this barrier, a number of common data models (CDMs) have been developed (e.g., Sentinel, the National Patient-Centered Clinical Research Network [PCORnet], Informatics for Integrating Biology in the Bedside [i2b2], the Observational Health Data Sciences and Informatics’ (OHDSI) Observational Medical Outcomes Partnership [OMOP]).[7] CDMs provide a standard set of terms (i.e., vocabulary) and structure so that data from different EMRs can be harmonized (Table 1, *Harmonized Data column*) and then aggregated.[8–10] Of these CDMs, the OMOP CDM[11, 12] has gained popularity due to its broad coverage of standard terminologies, flexibility and simplicity, and robust open-source tools.[9, 13–16]

**Table 1.** Demonstration of common data models to harmonize three commonly used assessments of function.

| Functional Assessment | Healthcare System 1 |  | Healthcare System 2 |  | Harmonized Data<sup>a</sup> |  |
| --- | --- | --- | --- | --- | --- | --- |
|  | Terminology | Location | Terminology | Location | Terminology | Location |
| **6-minute Walk Test** | 304700083 | derived_flowsheet | 3042800623 | clarity_flowsheet | 40766814 | Measurement |
| **AM-PAC Basic Mobility 6 Click** | 30441058001 | derived_flowsheet | 3042801216 | clarity_flowsheet | 4165295 | Measurement |

*Abbreviations: AM-PAC: Activity Measure for Post-Acute Care*
<sup>a</sup> *These are codes from the Observational Medical Outcomes Partnership Common Data Model (OMOP CDM); however, information in this table is intended to demonstrate the importance of data harmonization using common data models. As detailed in the manuscript and shown in the Supplemental Table, these are not the codes we are recommending for these functional measures.*

To leverage the OMOP CDM, healthcare systems must map their data into that CDM through an extract-transform-load (ETL) process. In this process, key data elements are first identified or extracted. These key data elements are then mapped into the OMOP CDM. Finally, the data from the EMR is loaded into the OMOP CDM. Thus, the utility of the OMOP CDM for creating robust datasets that can be used for high-value care is dependent on how well functional assessments are represented in the OMOP CDM. As a result, many medical fields have examined the extent to which key metrics in their field are included in the OMOP CDM.[14, 15, 17, 18] However, despite the importance of function to patients and to value-based care, the extent to which key functional assessments are included in the OMOP CDM has not been examined and, therefore, these important assessments are rarely included in large, real-world datasets.

## 2. OBJECTIVES

Our purpose was to determine the extent to which assessments of function that are recommended for use in neurologic and orthopaedic patient populations are contained within the OMOP CDM. These selected these two patient groups because they represent a large portion of patients seen in rehabilitation settings and of health care spending in the United States.[19, 20] We included assessments of function that are recommended in existing guidelines from the American Physical Therapy Association as physical therapy is a specific area of medicine that is focused on function. We hypothesized that 1) there would be good agreement between reviewers related to the mapping of the assessments and 2) that less than half of the included assessments would be included in the OMOP CDM. This work serves as a first step to ensuring that measures of function can be harmonized across healthcare systems and included in databases that facilitate our shift towards high value healthcare.

## 3. METHODS

This work was determined to not involve human subjects by the University of Utah IRB.

### 3.1. Assessment Selection

Because there are numerous assessments for function and quality of life in individuals with neurologic and orthopaedic conditions, we used clinical practice guidelines to select assessments with strong psychometric properties and with strong recommendations for use in these patient groups.

Two authors (H.H. and M.A.F.) with over 30 years of combined experience in neurologic rehabilitation selected the neurologic assessments for inclusion. They leveraged the American Physical Therapy Association Academy of Neurologic Physical Therapy’ Evidence Database to Guide Effectiveness (EDGE) documents for individuals with Multiple Sclerosis,[21] Parkinson Disease,[22] spinal cord injury,[23] stroke[24], traumatic brain injury,[25] and vestibular disorders[26] to guide the selection process. These documents provide recommendations regarding what functional assessments should be used in each specific patient population based on a thorough review of the assessments’ psychometric properties (i.e., validity, reliability) by content matter experts. The recommendations within each EDGE document are “highly recommended,” “recommended,” “unable to recommend,” or “not recommended.” They further make recommendations regarding which assessments entry level physical therapy practitioners should be familiar. Assessments that were 1) highly recommended or recommended in more than one neurologic patient population and 2) recommended for entry level physical therapy practitioners were included.

Two authors (A.T. and J.M.) with over 48 years of combined experience in orthopaedic rehabilitation selected the orthopaedic assessments for inclusion. Because there are no EDGE documents for orthopaedic conditions, they leveraged the American Physical Therapy Association Academy of Orthopaedic Physical Therapy’s Clinical Practice Guidelines (CPGs) for achilles tendinopathy, [27] ankle instability,[28] carpal tunnel syndrome,[29] heel pain, [30] ACL injury Prevention,[31] knee ligamentous instability,[32] meniscal and cartilage lesions,[33] anterior knee pain,[34] hamstring injury,[35] hip fracture,[36] hip osteoarthritis,[37] non-arthritic hip pain,[38] lateral elbow pain,[39] low back pain,[40, 41] concussion,[42] neck pain,[43] occupational injury,[44] pelvic girdle pain,[45] and adhesive capsulitis.[46] These CPGs provide recommendations on assessments for the targeted patient population based on the current best evidence. Recommendations are graded as “A - Strong Evidence,” “B – Moderate Evidence,” “C-Weak Evidence,” “D - Conflicting Evidence,” “E -Theoretical Foundation Evidence,” “F - Expert Opinion.” All recommendations graded A, B, or C were included in the current work except for concussion assessments. These were graded as “F-expert opinion” but were included as they were the only measures recommended for concussion assessment.

If an assessment was identified as both a neurologic and orthopaedic assessment, only one team of reviewers completed the mapping process described below. These assessments are identified in Appendix A.

### 3.2 Initial Mapping

After identifying the functional assessments, we followed a similar process to that of previous work to assess their content coverage in OMOP.[14, 15, 17] Two reviewers independently mapped the neurologic (M.A.F. and H.H.) and the orthopaedic (P.H. and A.T.) assessments into the OMOP CDM using Usagi.[47] USAGI is one of the many open source tools developed by OHDSI to support the use of the OMOP CDM. Usagi uses term similarity to provide an automated suggestion for the standard OMOP CDM vocabulary term, called a concept identification (ID) number, for each functional assessment.[47] Two reviewers independently reviewed the suggested concept ID number provided by Usagi to determine 1) if the concept ID number was correct and 2) if there were other concept ID numbers onto which the assessment should be mapped. Reviewers selected all concept ID numbers that were standard concepts, as opposed to non-standard, in the OMOP CDM that were appropriate for each assessment.

### 3.3 Metrics of Initial Agreement

After the initial mapping, each reviewer exported their mappings from Usagi for analysis. Two primary metrics were calculated separately for the neurologic assessments and for the orthopaedic assessments using R (version 4.2.1)[48]. The first metric was assessment agreement, in which each assessment was labeled as being 1) mapped by both reviewers, 2) mapped by reviewer A but not reviewer B, 3) mapped by reviewer B, but not reviewer A, or 4) unmapped by both reviewers. The second metric was the concept ID number agreement. This metric looked specifically at the assessments that were mapped by both reviewers to determine if the two reviewers mapped each assessment to the same concept ID number. Each concept ID number was categorized into one of three categories: 1) mapped by both reviewers, 2) mapped by reviewer A but not reviewer B, or 3) mapped by reviewer B, but not reviewer A. There was no unmapped category for this metric as we focused on only measures that were mapped by both reviewers to at least one concept ID number.

### 3.4 Reconciliation

After the analysis of the initial mapping, each reviewer team reviewed the following: 1) assessments that were unmapped by both reviewers, 2) assessments that were mapped by one but not the other reviewer, and 3) concept ID numbers that were mapped by one but not the other reviewer. The reviewer teams reconciled any differences in the mappings in these three areas. Following reconciliation, the same metrics described in the *Metrics of Initial Agreement* were calculated on the reconciled mappings.

### 3.5 Statistical Analysis

For assessment agreement and concept ID number agreement, we report 1) the portion of metrics in each category, 2) Cohen’s kappa,[49], 3) the percent overall agreement, and 4) Gwet’s agreement coefficient.[50, 51] We calculate Gwet’s agreement coefficient because there is a well-documented paradox of having a high percent overall agreement with a low kappa when the observations are imbalanced as expected in the analysis of concept ID number agreement (i.e., we expect no observations in the unmapped category).[50, 52–54] The Gwet’s agreement coefficient is interpreted similarly to Cohen’s kappa, with >0.81 being very good agreement, 0.61-0.8 being good agreement, 0.41 to 0.6 being moderate agreement, 0.21 to 0.4 being fair agreement, and less than or equal to 0.2 being poor agreement.[50] We also present Sankey plots to visualize changes in the mapping category from the initial mapping to reconciliation for each of these metrics. Lastly, we provide descriptive statistics of the number of concept ID numbers per assessment after reconciliation.

## 4. RESULTS

### 4.1 Assessment Agreement

We identified 160 unique assessments, 81 neurologic assessments and 79 orthopaedic assessments, to include in the analysis. During the initial mapping for the neurologic assessments, 48.1% (39/81) of assessments were mapped by both reviewers, 9.9% (8/81) were mapped by one but not the other reviewer, and 42% (34/81) were unmapped by both reviewers (Figure 1A). This resulted in a 90.1% overall agreement with a Cohen’s kappa and Gwet’s agreement coefficient of 0.80, indicating good agreement. After reconciliation, both reviewers mapped 46.9% of assessments (38/81), while the remaining 53.1% (43/81) were determined to be unmapped (Figure 1A). This resulted in a 100% overall agreement and a Cohen’s kappa and Gwet’s agreement coefficient of 1, indicating perfect agreement.

**Figure 1.**
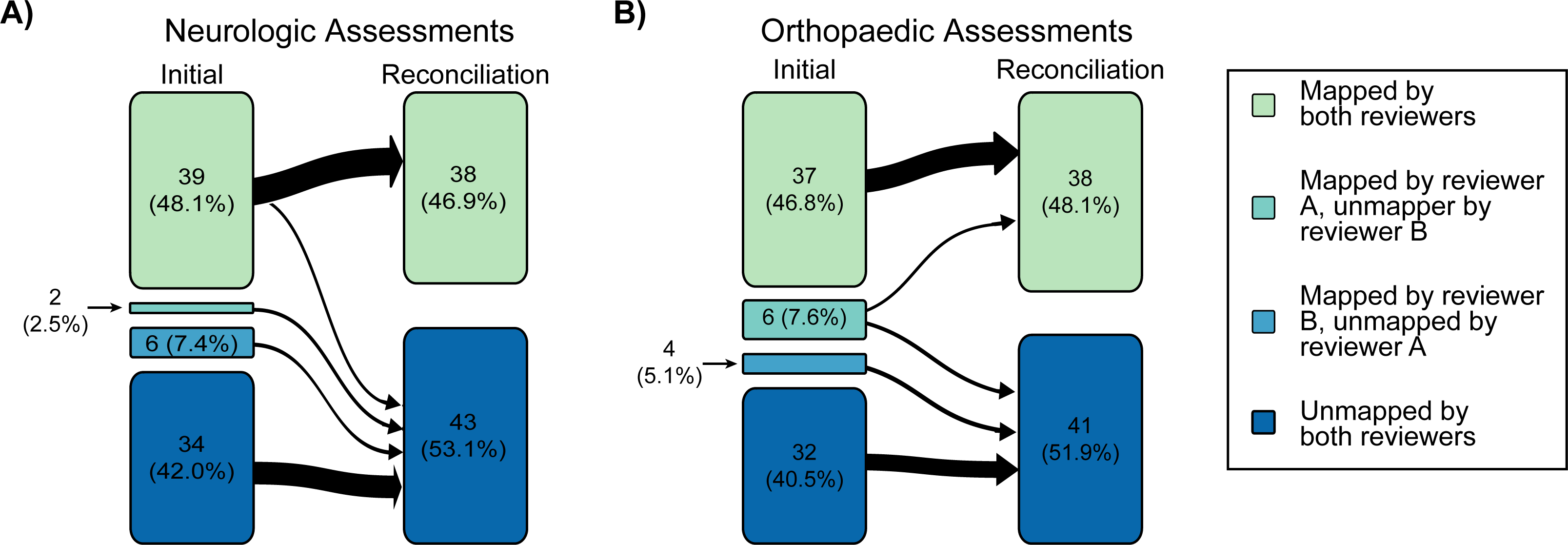
Sankey plot of (A) neurologic and (B) orthopaedic assessments after the initial mapping and after reconciliation.

During the initial mapping of the orthopaedic assessments, 46.8% (37/79) of the assessments were mapped by both reviewers, 12.7% (10/79) were mapped by one reviewer but not the other, and 48.1% (38/79) were unmapped by both reviewers (Figure 1B), This resulted in 87.3% overall agreement with a Cohen’s kappa and Gwet’s agreement coefficient of 0.74 and 0.75, respectively, indicating good agreement. After reconciliation, both reviewers mapped 48.1% of assessments (38/79), while the remaining 51.9% (41/79) were determined to be unmapped (Figure 1B). This resulted in a 100% overall agreement with a Cohen’s kappa and Gwet’s agreement coefficient of 1, indicating perfect agreement.

### 4.2 Concept ID number agreement

For the neurologic assessments that were initially mapped by both reviewers, there were 233 unique concept ID numbers identified. Of these, 33.5% (78/233) were the same between both reviewers, while 66.6% (155/233) were mapped by only one reviewer (Figure 2A). This resulted in a 33.5% overall agreement with Cohen’s kappa and Gwet’s agreement coefficient indicated poor agreement (-0.8 and -0.2, respectively). During reconciliation, it was determined that this poor agreement was in large part due to one reviewer selecting individual questionnaire items for several large testing batteries in addition to the full test. During reconciliation, these concept ID numbers were determined to be incorrect; thus, after completing the reconciliation process, 36.0% (84/233) concept ID numbers were determined to be correct, while 64.0% (149/233) were determined to be incorrect (Figure 2A). After reconciliation there was a 100% overall agreement with a Cohen’s kappa and Gwet’s agreement coefficient of 1, indicating perfect agreement. For the 38 neurologic assessments that were mapped, there were 84 unique concept ID numbers. Ten neurologic assessments had a single unique concept ID number with an average number of concept ID numbers per neurologic assessment of 2.2 (1.3) and a median of 2 (Figure 3A). Importantly, a number of assessments had concept ID numbers in multiple OMOP domains, typically in the measurement and observation domain (Appendix A).

**Figure 2.**
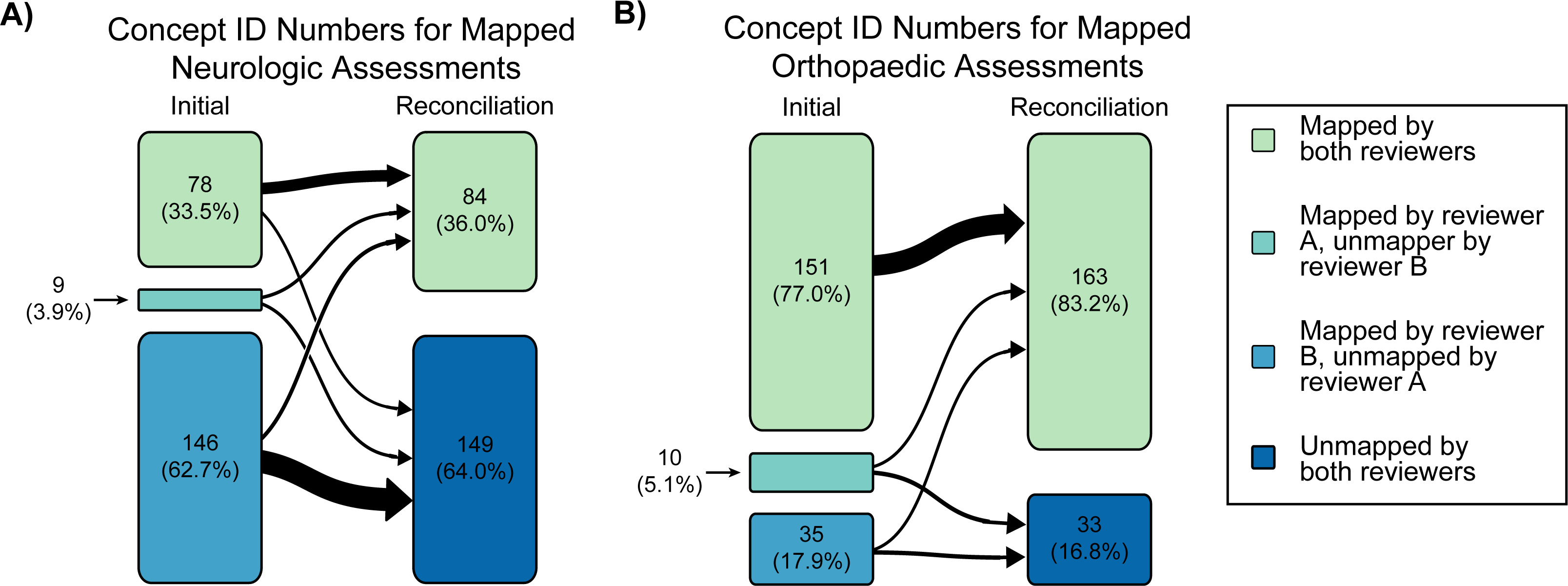
Sankey plots of (A) neurologic and (B) orthopaedic concept ID numbers after the initial mapping and after reconciliation.

**Figure 3.**
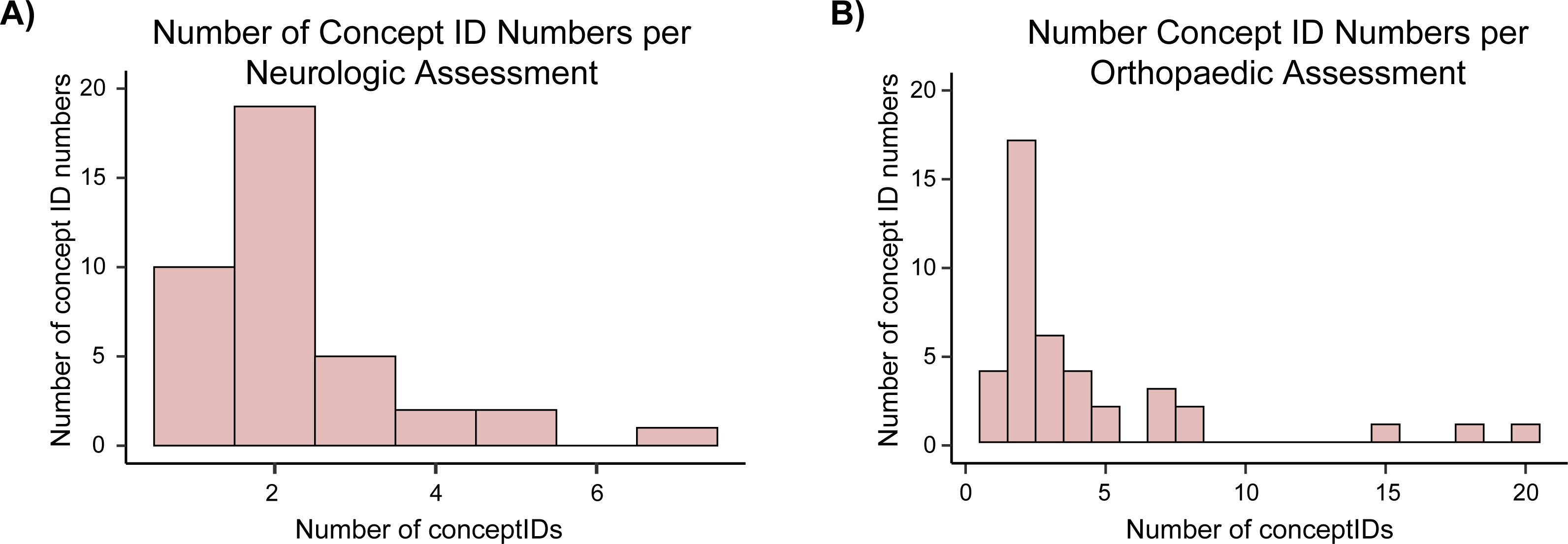
Number of concept ID numbers per (A) neurologic and (B) orthopaedic assessments that were mapped by both reviewers after reconciliation.

For the orthopaedic assessments that were initially mapped, there were 196 unique concept ID numbers identified. Of these, 77.0% (151/196) were the same between both reviewers, while 23.0% (45/196) were mapped by only one reviewer (Figure 2B). The overall percent agreement was 77.0%. Cohen’s kappa and Gwet’s agreement coefficient were -0.09 and 0.71, respectively. This demonstrates the well-documented paradox that can occur with Cohen’s kappa when observations are imbalanced, [50, 52–54] necessitating the inclusion of Gwet’s agreement.[51] After reconciliation, 83.2% (163/196) concept ID numbers were determined to be correct, while 16.8% (33/196) were determined to be incorrect (Figure 2B). After reconciliation, overall percent agreement was 100% with a Cohen’s kappa and Gwet’s agreement coefficient of 1, indicating perfect agreement. The final mapping for orthopaedic assessments resulted in 163 unique concept ID numbers for the 38 orthopaedic measures that were mapped. Only four orthopaedic assessments had a single unique concept ID number. The average number of concept ID numbers per orthopaedic assessment was 4.3 (4.4) with a median of 2.5 (Figure 3B). Many of the assessments with a large number of potential concept ID numbers were the Patient-Reported Outcomes Measurement Information System (PROMIS) measures as there are numerous versions of these assessments in the OMOP CDM (Appendix A). As with the neurologic assessments, many measures had at least one concept ID number in the measurement and observation domain of the OMOP CDM.

## 5. DISCUSSION

This study examined the extent to which functional assessments are included in the OMOP CDM specifically for neurologic and orthopaedic conditions. Our hypotheses were supported such that there was high agreement between reviewers and that less than 50% of assessments were included in the OMOP CDM. We also found that there were multiple standard OMOP CDM concept ID numbers, or vocabulary terms, for most assessments.

We found that 46.9% and 48.1% of the neurologic and orthopaedic assessments were included in the OMOP CDM after reconciliation. The optimistic conclusion from this finding is that almost half of the assessments that are recommended to measure function in these two broad patient groups are already in the OMOP CDM; thus, because of the importance of these constructs to all patients, these assessments should be included in datasets that leverage OMOP CDM. Of note, although we selected these assessments based on recommendations for neurologic and orthopaedic conditions, many of the assessments are valid in other patient populations and, therefore, can be used across an even broad group of patients. For example, the European Quality of Life was included as an orthopaedic assessment. This assessment, however, has been validated in neurologic[55, 56] and general populations[57] as well. Similarly, the 10 meter walk test, which is used to calculate gait speed, was included as a neurologic assessment; however, this metric has been called the “sixth vital sign”[58] and is a critical metric across all patient diagnoses. This highlights that the mapping of functional assessments has significance across medical diagnoses and that the OMOP CDM supports the harmonization of some of these assessments.

The more pessimistic interpretation of our findings is that over half of the assessments we selected based on clinical practice guidelines and recommendations from the American Association for Physical Therapy are not included in the OMOP CDM. Some of the assessments not included in the OMOP CDM are particularly troublesome because of their importance in guiding treatment and understanding patient outcomes. For example, the Functional Gait Assessment is a predictor of falls in neurologic patient groups[59–61] and, is a recommended functional assessment in all individuals with neurologic diagnoses.[62] This tool is also reliable, valid, and predictive of falls in geriatric populations.[63, 64] Yet, this assessment is not mapped into the OMOP CDM. Similarly, the Fear Avoidance Beliefs Questionnaire is recommended with strong evidence in 3 CPGs[41, 44, 65] and with weak evidence in 1 other CPG.[28] Further, fear avoidance is known to significantly affect quality of life and function,[66, 67] yet it is not mapped into the OMOP CDM. Our findings and these specific examples demonstrate the need for expanding the OMOP CDM vocabulary to include more assessments of function.

A significant limitation in the coverage of functional assessments in the OMOP CDM that we identified is that many assessments had multiple standard concept ID numbers. This lack of unique concept ID number is problematic as individuals undertaking an ETL at one site may select a different concept ID number than the individual conducting the ETL process at a different site. This challenge limits our ability to harmonize these assessments across healthcare systems with the OMOP CDM. Agreement and guidelines about which concept ID number should be used from functional assessments are needed. Providing these recommendations is beyond the scope of this work and will require collaboration across the OHDSI community. Based on this work, there are two primary areas that these guidelines should address. First, there should be clarification regarding onto which domain of the OMOP CDM the functional assessment should be mapped. This clarification is needed because many assessments had a single unique concept ID number when constrained to a single domain of the OMOP CDM. For example, we identified two appropriate concept ID numbers for the Short Physical Performance Battery, which is a commonly used functional assessment in orthopaedic and geriatric patient groups; one of the concept ID numbers was in the measurement domain, while the other was in the observation domain (Appendix A). This situation occurred in a number of assessments; thus, clarification on which domain is correct would help mitigate the challenge with multiple concept ID numbers. Secondly, guidelines should address assessments that have multiple concept ID numbers within the same domain. For example, the 6-minute walk test is a commonly used assessment of walking endurance and has three vocabulary terminologies in the OMOP CDM (Appendix A). This could be address by providing clear recommendations for which concept ID number to use or by revising the OMOP CDM vocabulary to minimize redundancy within the CDM.

While the current work significantly contributes to our ability to use the OMOP CDM, there are several limitations. First, we focused on two broad patient diagnosis categories: neurologic and orthopaedic. While these patient groups make up a significant portion of healthcare spending, there are numerous other diagnoses for which a similar exploration should be performed. Importantly, however, many of the assessments included in this analysis can be and are used in other patient populations. Additionally, there are numerous assessments of function that we did not include in our analysis. Instead, we used clinical practice guidelines and other recommendations to guide the selection of the assessments we included, yet there may be other assessments of function that researchers and clinical teams may want to map into the OMOP CDM. Finally, we focused primarily on physical function in this work; however, other domains of function, such as cognition and mood, are important to patients and to value-based care initiatives. Therefore, similar work with these other domains of function would be valuable.

## 6. CONCLUSION

In this work, we found that the OMOP CDM includes a portion of functional assessments that are recommended for use in clinical practice with individuals with neurologic or orthopaedic conditions. While these findings are encouraging, many of the assessments that were mapped did not have a single unique term in the OMOP CDM. To include functional assessments in databases that allow us to understand and improve the value of healthcare, we must 1) ensure that the assessments that are already mapped into the OMOP CDM are included in the development of large databases, 2) work towards guidelines for the ETL process of functional assessments into the OMOP CDM terminologies, and 3) continuing to expand the OMOP CDM such that it includes all key functional assessments.

## Data Availability

All data produced in this work are included in the manuscript.

## APPENDICES

Appendix A

## REFERENCES

1. Papanicolas I, Woskie LR, Jha AK. Health Care Spending in the United States and Other High-Income Countries. Jama 2018; 319: 1024–1039. doi:10.1001/jama.2018.1150

2. Munira Z. Gunja, Evan D. Gumas, II Rdw. U.S. Health Care from a Global Perspective, 2022: Accelerating Spending, Worsening Outcomes. Commonwealth Fund 2023. 10.26099/8ejy-yc74

3. Jewell DV, Moore JD, Goldstein MS. Delivering the physical therapy value proposition: a call to action. Phys Ther 2013; 93: 104–114. doi:10.2522/ptj.20120175

4. Porter ME. What is value in health care? N Engl J Med 2010; 363: 2477–2481. doi:10.1056/NEJMp1011024

5. Teisberg E, Wallace S, O’Hara S. Defining and Implementing Value-Based Health Care: A Strategic Framework. Acad Med 2020; 95: 682–685. doi:10.1097/acm.0000000000003122

6. Baumhauer JF, Bozic KJ. Value-based Healthcare: Patient-reported Outcomes in Clinical Decision Making. Clin Orthop Relat Res 2016; 474: 1375–1378. doi:10.1007/s11999-016-4813-4

7. Weeks J, Pardee R. Learning to Share Health Care Data: A Brief Timeline of Influential Common Data Models and Distributed Health Data Networks in U.S. Health Care Research. EGEMS (Wash DC) 2019; 7: 4. doi:10.5334/egems.279

8. Voss EA, Makadia R, Matcho A et al. Feasibility and utility of applications of the common data model to multiple, disparate observational health databases. J Am Med Inform Assoc 2015; 22: 553–564. doi:10.1093/jamia/ocu023

9. Garza M, Del Fiol G, Tenenbaum J et al. Evaluating common data models for use with a longitudinal community registry. J Biomed Inform 2016; 64: 333–341. doi:10.1016/j.jbi.2016.10.016

10. Kent S, Burn E, Dawoud D, et al. Common Problems, Common Data Model Solutions: Evidence Generation for Health Technology Assessment. Pharmacoeconomics 2021; 39: 275–285. doi:10.1007/s40273-020-00981-9

11. Hripcsak G, Duke JD, Shah NH et al. Observational Health Data Sciences and Informatics (OHDSI): Opportunities for Observational Researchers. Stud Health Technol Inform 2015; 216: 574–578.

12. Observational Health Data Sciences and Informatics. The Book of OHDSI Available at: https://ohdsi.github.io/TheBookOfOhdsi/; Date Accessed: February 23, 2024.

13. Klann JG, Joss MAH, Embree K et al. Data model harmonization for the All Of Us Research Program: Transforming i2b2 data into the OMOP common data model. PLoS One 2019; 14: e0212463. doi:10.1371/journal.pone.0212463

14. Cho S, Sin M, Tsapepas D et al. Content Coverage Evaluation of the OMOP Vocabulary on the Transplant Domain Focusing on Concepts Relevant for Kidney Transplant Outcomes Analysis. Appl Clin Inform 2020; 11: 650–658. doi:10.1055/s-0040-1716528

15. Sathappan SMK, Jeon YS, Dang TK et al. Transformation of Electronic Health Records and Questionnaire Data to OMOP CDM: A Feasibility Study Using SG_T2DM Dataset. Appl Clin Inform 2021; 12: 757–767. doi:10.1055/s-0041-1732301

16. Reinecke I, Zoch M, Reich C et al. The Usage of OHDSI OMOP - A Scoping Review. Stud Health Technol Inform 2021; 283: 95–103. doi:10.3233/shti210546

17. Cai CX, Halfpenny W, Boland MV et al. Advancing Toward a Common Data Model in Ophthalmology: Gap Analysis of General Eye Examination Concepts to Standard Observational Medical Outcomes Partnership (OMOP) Concepts. Ophthalmol Sci 2023; 3: 100391. doi:10.1016/j.xops.2023.100391

18. Lamer A, Abou-Arab O, Bourgeois A et al. Transforming Anesthesia Data Into the Observational Medical Outcomes Partnership Common Data Model: Development and Usability Study. J Med Internet Res 2021; 23: e29259. doi:10.2196/29259

19. Dieleman JL, Cao J, Chapin A et al. US Health Care Spending by Payer and Health Condition, 1996-2016. JAMA 2020; 323: 863–884. doi:10.1001/jama.2020.0734

20. Feigin VL, Forouzanfar MH, Krishnamurthi R et al. Global and regional burden of stroke during 1990-2010: findings from the Global Burden of Disease Study 2010. Lancet 2014; 383: 245–254. doi:10.1016/s0140-6736(13)61953-4

21. Potter K, Cohen ET, Allen DD et al. Outcome measures for individuals with multiple sclerosis: recommendations from the American Physical Therapy Association Neurology Section task force. Phys Ther 2014; 94: 593–608. doi:10.2522/ptj.20130149

22. Academy of Neurologic Physical Therapy. Parkinson Evidence Database to Guide Effectiveness. In: 2018:

23. Kahn JH, Tappan R, Newman CP et al. Outcome Measure Recommendations From the Spinal Cord Injury EDGE Task Force. Phys Ther 2016; 96: 1832–1842. doi:10.2522/ptj.20150453

24. Sullivan JE, Crowner BE, Kluding PM et al. Outcome measures for individuals with stroke: process and recommendations from the American Physical Therapy Association neurology section task force. Phys Ther 2013; 93: 1383–1396. doi:10.2522/ptj.20120492

25. McCulloch KL, de Joya AL, Hays K et al. Outcome Measures for Persons With Moderate to Severe Traumatic Brain Injury: Recommendations From the American Physical Therapy Association Academy of Neurologic Physical Therapy TBI EDGE Task Force. J Neurol Phys Ther 2016; 40: 269–280. doi:10.1097/npt.0000000000000145

26. Academy of Neurologic Physical Therapy. Vestibular EDGE. In: 2018:

27. Martin RL, Chimenti R, Cuddeford T et al. Achilles Pain, Stiffness, and Muscle Power Deficits: Midportion Achilles Tendinopathy Revision 2018. J Orthop Sports Phys Ther 2018; 48: A1–a38. doi:10.2519/jospt.2018.0302

28. Martin RL, Davenport TE, Fraser JJ et al. Ankle Stability and Movement Coordination Impairments: Lateral Ankle Ligament Sprains Revision 2021. J Orthop Sports Phys Ther 2021; 51: Cpg1–cpg80. doi:10.2519/jospt.2021.0302

29. Erickson M, Lawrence M, Jansen CWS et al. Hand Pain and Sensory Deficits: Carpal Tunnel Syndrome. J Orthop Sports Phys Ther 2019; 49: Cpg1–cpg85. doi:10.2519/jospt.2019.0301

30. Koc TA, Jr., Bise CG, Neville C et al. Heel Pain - Plantar Fasciitis: Revision 2023. J Orthop Sports Phys Ther 2023; 53: Cpg1–cpg39. doi:10.2519/jospt.2023.0303

31. Arundale AJH, Bizzini M, Dix C et al. Exercise-Based Knee and Anterior Cruciate Ligament Injury Prevention. J Orthop Sports Phys Ther 2023; 53: Cpg1–cpg34. doi:10.2519/jospt.2023.0301

32. Logerstedt DS, Snyder-Mackler L, Ritter RC et al. Knee stability and movement coordination impairments: knee ligament sprain. J Orthop Sports Phys Ther 2010; 40: A1–a37. doi:10.2519/jospt.2010.0303

33. Logerstedt DS, Scalzitti DA, Bennell KL et al. Knee Pain and Mobility Impairments: Meniscal and Articular Cartilage Lesions Revision 2018. J Orthop Sports Phys Ther 2018; 48: A1–a50. doi:10.2519/jospt.2018.0301

34. Willy RW, Hoglund LT, Barton CJ, et al. Patellofemoral Pain. J Orthop Sports Phys Ther 2019; 49: Cpg1-cpg95. doi:10.2519/jospt.2019.0302

35. Martin RL, Cibulka MT, Bolgla LA et al. Hamstring Strain Injury in Athletes. J Orthop Sports Phys Ther 2022; 52: Cpg1–cpg44. doi:10.2519/jospt.2022.0301

36. McDonough CM, Harris-Hayes M, Kristensen MT et al. Physical Therapy Management of Older Adults With Hip Fracture. J Orthop Sports Phys Ther 2021; 51: Cpg1–cpg81. doi:10.2519/jospt.2021.0301

37. Cibulka MT, Bloom NJ, Enseki KR, et al. Hip Pain and Mobility Deficits-Hip Osteoarthritis: Revision 2017. J Orthop Sports Phys Ther 2017; 47: A1-a37. doi:10.2519/jospt.2017.0301

38. Enseki KR, Bloom NJ, Harris-Hayes M et al. Hip Pain and Movement Dysfunction Associated With Nonarthritic Hip Joint Pain: A Revision. J Orthop Sports Phys Ther 2023; 53: Cpg1–cpg70. doi:10.2519/jospt.2023.0302

39. Lucado AM, Day JM, Vincent JI, et al. Lateral Elbow Pain and Muscle Function Impairments. J Orthop Sports Phys Ther 2022; 52: Cpg1-cpg111. doi:10.2519/jospt.2022.0302

40. Delitto A, George SZ, Van Dillen L, et al. Low back pain. J Orthop Sports Phys Ther 2012; 42: A1-57. doi:10.2519/jospt.2012.42.4.A1

41. George SZ, Fritz JM, Silfies SP et al. Interventions for the Management of Acute and Chronic Low Back Pain: Revision 2021. J Orthop Sports Phys Ther 2021; 51: Cpg1–cpg60. doi:10.2519/jospt.2021.0304

42. Quatman-Yates CC, Hunter-Giordano A, Shimamura KK et al. Physical Therapy Evaluation and Treatment After Concussion/Mild Traumatic Brain Injury. J Orthop Sports Phys Ther 2020; 50: Cpg1–cpg73. doi:10.2519/jospt.2020.0301

43. Blanpied PR, Gross AR, Elliott JM, et al. Neck Pain: Revision 2017. J Orthop Sports Phys Ther 2017; 47: A1-a83. doi:10.2519/jospt.2017.0302

44. Daley D, Payne LP, Galper J et al. Clinical Guidance to Optimize Work Participation After Injury or Illness: The Role of Physical Therapists. J Orthop Sports Phys Ther 2021; 51: Cpg1–cpg102. doi:10.2519/jospt.2021.0303

45. Clinton S, Newell A, Downey P et al. Pelvic Girdle Pain in the Antepartum Population: Physical Therapy Clinical Practice Guidelines Linked to the International Classification of Functioning, Disability, and Health From the Section on Women’s Health and the Orthopaedic Section of the American Physical Therapy Association. Journal of Women_Js Health Physical Therapy 2017; 41: 102–125. doi:10.1097/JWH.0000000000000081

46. Kelley MJ, Shaffer MA, Kuhn JE et al. Shoulder pain and mobility deficits: adhesive capsulitis. J Orthop Sports Phys Ther 2013; 43: A1–31. doi:10.2519/jospt.2013.0302

47. Usagi. OHDSI Usagi Available at: https://ohdsi.github.io/Usagi/; Date Accessed: February 23, 2024.

48. Team RC. R: A language and environment for statistical computing. R Foundation for Statistical Computing, Vienna, Austria 2022.

49. McHugh ML. Interrater reliability: the kappa statistic. Biochem Med (Zagreb) 2012; 22: 276–282.

50. Dettori JR, Norvell DC. Kappa and Beyond: Is There Agreement? Global Spine J 2020; 10: 499–501. doi:10.1177/2192568220911648

51. Gwet KL. Computing inter-rater reliability and its variance in the presence of high agreement. Br J Math Stat Psychol 2008; 61: 29–48. doi:10.1348/000711006×126600

52. Cicchetti DV, Feinstein AR. High agreement but low kappa: II. Resolving the paradoxes. J Clin Epidemiol 1990; 43: 551–558. doi:10.1016/0895-4356(90)90159-m

53. Feinstein AR, Cicchetti DV. High agreement but low kappa: I. The problems of two paradoxes. J Clin Epidemiol 1990; 43: 543–549. doi:10.1016/0895-4356(90)90158-l

54. Viera AJ, Garrett JM. Understanding interobserver agreement: the kappa statistic. Fam Med 2005; 37: 360–363.

55. Chen P, Lin KC, Liing RJ et al. Validity, responsiveness, and minimal clinically important difference of EQ-5D-5L in stroke patients undergoing rehabilitation. Qual Life Res 2016; 25: 1585–1596. doi:10.1007/s11136-015-1196-z

56. van Agt HM, Essink-Bot ML, Krabbe PF et al. Test-retest reliability of health state valuations collected with the EuroQol questionnaire. Soc Sci Med 1994; 39: 1537–1544. doi:10.1016/0277-9536(94)90005-1

57. Brazier J, Jones N, Kind P. Testing the validity of the Euroqol and comparing it with the SF-36 health survey questionnaire. Qual Life Res 1993; 2: 169–180. doi:10.1007/bf00435221

58. Fritz S, Lusardi M. White paper: “walking speed: the sixth vital sign”. J Geriatr Phys Ther 2009; 32: 46–49.

59. Foreman KB, Addison O, Kim HS et al. Testing balance and fall risk in persons with Parkinson disease, an argument for ecologically valid testing. Parkinsonism Relat Disord 2011; 17: 166–171. doi:10.1016/j.parkreldis.2010.12.007

60. Lin JH, Hsu MJ, Hsu HW et al. Psychometric comparisons of 3 functional ambulation measures for patients with stroke. Stroke 2010; 41: 2021–2025. doi:10.1161/strokeaha.110.589739

61. Marchetti GF, Lin CC, Alghadir A et al. Responsiveness and minimal detectable change of the dynamic gait index and functional gait index in persons with balance and vestibular disorders. J Neurol Phys Ther 2014; 38: 119–124. doi:10.1097/npt.0000000000000015

62. Moore JL, Potter K, Blankshain K et al. A Core Set of Outcome Measures for Adults With Neurologic Conditions Undergoing Rehabilitation: A CLINICAL PRACTICE GUIDELINE. J Neurol Phys Ther 2018; 42: 174–220. doi:10.1097/npt.0000000000000229

63. Beninato M, Fernandes A, Plummer LS. Minimal clinically important difference of the functional gait assessment in older adults. Phys Ther 2014; 94: 1594–1603. doi:10.2522/ptj.20130596

64. Wrisley DM, Kumar NA. Functional gait assessment: concurrent, discriminative, and predictive validity in community-dwelling older adults. Phys Ther 2010; 90: 761–773. doi:10.2522/ptj.20090069

65. Simonds AH, Abraham K, Spitznagle T. Clinical Practice Guidelines for Pelvic Girdle Pain in the Postpartum Population. The Journal of Women’s & Pelvic Health Physical Therapy 2022; 46: E1–E38. doi:10.1097/jwh.0000000000000236

66. Baez SE, Hoch MC, Hoch JM. Psychological factors are associated with return to pre-injury levels of sport and physical activity after ACL reconstruction. Knee Surg Sports Traumatol Arthrosc 2020; 28: 495–501. doi:10.1007/s00167-019-05696-9

67. Burgess R, Mansell G, Bishop A et al. Predictors of functional outcome in musculoskeletal healthcare: An umbrella review. Eur J Pain 2020; 24: 51–70. doi:10.1002/ejp.1483

