## Appendix A for "Examination of the coverage of functional assessments in the OMOP common data model"

**Table.** Mapping of neurologic and orthopaedic rehabilitation clinical assessments to OMOP Standard concept IDs. Of note, there are multiple standard OMOP concept IDs for most measures. We are not recommending which one the rehabilitation community should use as this decision should be made by the broader OMOP community.

| **Assessment** | **Category^a^** | **OMOP Standard Concept Name** | **OMOP Standard Concept ID** | **OMOP Domain** |
| --- | --- | --- | --- | --- |
| **10 Meter Walk Test** | Neurologic | 4-Meter Walk Gait Speed Test [NIH Toolbox] | 42529267 | Measurement |
|  |  | Timed six meter walk time | 3655978 | Observation |
|  |  | Timed six meter walk gait speed | 3655982 |  |
|  |  | Gait speed | 37110142 |  |
|  |  | Timed 10 meter walk | 4197638 | Procedure |
| **12-Item Multiple Sclerosis Walking Scale** | Neurologic | Unmapped | | |
| **2 Minute Walk** | Neurologic | Walk distance score 2 minutes [NIH Toolbox] | 42529361 | Measurement |
|  |  | 2-Minute Walk Endurance Test [NIH Toolbox] | 42529266 |  |
| **30 Second Chair Stand** | Neurologic | Thirty second chair stand test | 42872744 | Measurement |
|  |  | Thirty second chair stand test score | 42872746 | Observation |
|  |  | Sit to stand frequency in 30 seconds | 40768924 |  |
| **5 Times Sit to Stand** | Neurologic | Unmapped | | |
| **6 Minute Walk Test** | Neurologic | 6 minute walk test distance | 606289 | Measurement |
|  |  | 6 minute walk test distance | 44806532 |  |
|  |  | Six minute walk test | 40766814 |  |
| **9 Hole Peg Test** | Neurologic | Nine hole peg test | 4165291 | Measurement |
|  |  | 9-Hole Pegboard Dexterity Test Time Hand - left [NIH Toolbox] | 42529303 |  |
|  |  | 9-Hole Pegboard Dexterity Test Time Hand - right [NIH Toolbox] | 42529304 |  |
|  |  | Nine hole peg test score | 40488387 | Observation |
| **Action Research Arm Test** | Neurologic | Action research arm test | 36684870 | Measurement |
| **Activities-Specific Balance Confidence Scale** | Neurologic | Activities specific balance confidence scale | 40481019 | Measurement |
|  |  | Activities specific balance confidence score | 40480116 | Observation |
| **Activity Measure for Post Acute Care Mobility** | Neurologic | Basic mobility score [AM-PAC] | 21491085 | Measurement |
|  |  | Basic mobility items number [Activity Measure for Post-Acute Care] | 21491087 | Observation |
| **Agitated Behavior Scale** | Neurologic | Unmapped | | |
| **Anterior Cruciate Ligament-Return to Sport After Injury (ACL-RSI)** | Orthopaedic | Unmapped | | |
| **Anterior Knee Pain Scale** | Orthopaedic | Unmapped | | |
| **ASIA Impairment Scale** | Neurologic | ASIA impairment scale | 1616330 | Measurement |
| **Assessment of Life Habits** | Neurologic | Unmapped | | |
| **Back Disability Risk Questionnaire** | Orthopaedic | Unmapped | | |
| **Balance Error Scoring System** | Neurologic | Modified Balance Error Scoring System | 36675001 | Measurement |
| **Balance Evaluation Systems Test (BESTest)** | Neurologic | Unmapped | | |
| **Berg Balance Scale** | Neurologic | Berg balance scale | 4326493 | Measurement |
|  |  | Berg balance test | 4232747 |  |
|  |  | Berg balance score | 44802297 | Observation |
| **Boston Carpal Tunnel Questionnaire Functional Scale** | Orthopaedic | Unmapped | | |
| **Boston Carpal Tunnel Questionnaire Symptom Severity Scale** | Orthopaedic | Unmapped | | |
| **Box and Block Test** | Neurologic | Unmapped | | |
| **Brief Balance Evaluation Systems Test (Brief BESTest)** | Neurologic | Unmapped | | |
| **Canadian Occupational Performance Measure** | Neurologic | Canadian occupational performance measure | 4231508 | Measurement |
|  |  | Canadian occupational performance measure score | 40481486 | Observation |
| **Clinical Test of Sensory Interaction and Balance** | Neurologic | Pediatric clinical test of sensory integration and balance | 4165297 | Measurement |
| **Coma Recovery Scale- Revised** | Neurologic | Unmapped | | |
| **Community Integration Questionnaire** | Neurologic | Unmapped | | |
| **Community Mobility and Balance Scale** | Neurologic | Unmapped | | |
| **Craig Handicap Assessment and Reporting Technique** | Neurologic | Unmapped | | |
| **Craig Hospital Inventory of Environmental Factors** | Neurologic | Unmapped | | |
| **Cranial Cervical Flexion Test** | Orthopaedic | Unmapped | | |
| **Cumberland Ankle Instability Tool (Dx)** | Orthopaedic | Unmapped | | |
| **Cumulated Ambulation Score** | Orthopaedic | Ambulate | 45880566 | Meas Value |
| **Deep Neck Flexor Endurance Test** | Orthopaedic | Unmapped | | |
| **Dellon-Modified Moberg Pick-Up Test** | Orthopaedic | Moberg pickup test | 40485480 | Measurement |
|  |  | Moberg pickup test score | 40491928 | Observation |
| **Disabilities of the Arm, Shoulder And Hand (Dash)** | Orthopaedic | Disabilities of the arm, shoulder and hand questionnaire | 601842 | Measurement |
|  |  | Disabilities of the arm shoulder and hand questionnaire | 40481154 |  |
|  |  | Disabilities of the arm shoulder and hand outcome measure score | 40481564 |  |
|  |  | Disabilities of the arm shoulder and hand outcome measurement sports/performing arts module score | 44782454 |  |
|  |  | Disabilities of the arm shoulder and hand outcome measure work module score | 45772884 |  |
|  |  | Quick disabilities of the arm shoulder and hand outcome measure score | 40492187 |  |
|  |  | Quick disabilities of the arm shoulder and hand outcome measurement | 44813074 |  |
| **Disability Rating Index (DRI)** | Orthopaedic | Unmapped | | |
| **Disability Rating Scale** | Neurologic | Disability rating scale | 4167758 | Measurement |
|  |  | Disability Rating Scale score | 36713777 | Observation |
| **Disease Steps** | Neurologic | Unmapped | | |
| **Dix Hallpike** | Neurologic | Dix-Hallpike maneuver | 4138451 | Procedure |
| **Dizziness Handicap Inventory (DHI) ^b^** | Neurologic | Dizziness Handicap Inventory | 46284659 | Measurement |
|  |  | Dizziness Handicap Inventory score | 36714213 | Observation |
| **Dynamic Gait Index (DGI)** | Neurologic | Dynamic gait index | 40481604 | Measurement |
|  |  | Dynamic gait index score | 40482816 | Observation |
| **Dynamic Visual Acuity Testing (DVAT)** | Orthopaedic | Unmapped | | |
| **European Quality of Life 5-Dimensions ^b^** | Orthopaedic | EuroQol five dimension three level scale | 701333 | Measurement |
|  |  | EuroQoL five dimension questionnaire | 40483273 |  |
|  |  | EuroQol five dimension five level scale | 44806410 |  |
|  |  | EuroQol five dimension self-report questionnaire | 44807984 |  |
|  |  | EuroQol five dimension three level questionnaire | 600634 |  |
|  |  | EuroQol five dimension five level index value | 42537273 | Observation |
|  |  | EuroQol five dimension self-report questionnaire score | 44807986 |  |
| **Falls Efficacy Scale-International** | Orthopaedic | FES-I - Falls Efficacy Scale International | 3657176 | Measurement |
|  |  | Short falls efficacy scale - international | 40480721 |  |
|  |  | FES-I (Falls Efficacy Scale International) score | 3657177 | Observation |
|  |  | Short falls efficacy scale - international score | 40481994 |  |
| **Fatigue Scale for Motor and Cognitive Functions** | Neurologic | Unmapped | | |
| **Fear Avoidance Behavioral Questionnaire** | Orthopaedic | Unmapped | | |
| **Foot And Ankle Ability Measure (FAAM)** | Orthopaedic | Unmapped | | |
| **Foot Function Index (FFI)** | Orthopaedic | Foot Function Index | 35621854 | Measurement |
|  |  | Foot Function Index total score | 35621856 | Observation |
| **Foot Health Status Questionnaire (FHSQ)** | Orthopaedic | Unmapped | | |
| **Foot Posture Index-6 (FPI-6)** | Orthopaedic | Foot Posture Index | 35609129 | Measurement |
|  |  | Foot Posture Index score | 35609170 | Observation |
| **Four Square Step Test** | Neurologic | Four step square test | 36684828 | Measurement |
| **Freezing Of Gait Questionnaire** | Neurologic | Unmapped | | |
| **Fugl Meyer Assessment of Motor Performance** | Neurologic | Unmapped | | |
| **Functional Assessment for Acute HS Injuries** | Orthopaedic | Unmapped | | |
| **Functional Assessment of Multiple Sclerosis** | Neurologic | Unmapped | | |
| **Functional Capacity Evaluation** | Orthopaedic | Functional capacity evaluation | 40481051 | Measurement |
|  |  | Functional capacity evaluation score | 40486509 | Observation |
| **Functional Gait Assessment** | Neurologic | Unmapped | | |
| **Functional Independence Measure ^b^** | Neurologic | Functional independence measure | 4159704 | Measurement |
|  |  | Functional Independence Measure score | 37396842 | Observation |
| **Functional Reach Test** | Neurologic | Functional reach | 4232748 | Observation |
|  |  | Functional reach test | 4323348 | Procedure |
| **Functional Status Exam** | Neurologic | Unmapped | | |
| **Glasgow Coma Scale** | Neurologic | Glasgow coma scale | 3032652 | Measurement |
|  |  | Glasgow coma scale | 4296538 |  |
|  |  | Glasgow coma score total | 3007194 |  |
|  |  | Glasgow coma score | 4093836 | Observation |
| **Harris Hip Score** | Orthopaedic | Harris hip score scale | 40481155 | Measurement |
|  |  | Harris hip score | 40481979 | Observation |
|  |  | Total score [Harris Hip Score] | 1761583 |  |
|  |  | Harris Hip Score panel [Harris Hip Score] | 1761680 |  |
| **Head Impulse Test** | Neurologic | Head impulse test | 36715427 | Procedure |
| **High Level Mobility Assessment ^b^** | Neurologic | Unmapped | | |
| **Impact on Participation and Autonomy Questionnaire** | Neurologic | Unmapped | | |
| **International Knee Documentation Committee (IKDC 2000)** | Orthopaedic | Unmapped | | |
| **Jebsen Taylor Arm Function Test** | Neurologic | Jebsen hand function test | 4169153 | Measurement |
|  |  | Jebsen-Taylor hand function test score | 40486014 | Observation |
| **Knee Injury and Osteoarthritis Outcome Score (KOOS)** | Orthopaedic | Knee Injury and Osteoarthritis Outcome Score | 37203804 | Measurement |
|  |  | Knee injury and Osteoarthritis Outcome Score [KOOS] | 42869846 | Observation |
| **Knee Injury and Osteoarthritis Outcome Score: Patellofemoral Subscale (KOOS-PF)** | Orthopaedic | Unmapped | | |
| **Knee Quality of Life 26-Item** | Orthopaedic | Unmapped | | |
| **Limb Symmetry Index** | Orthopaedic | Unmapped | | |
| **Lower Extremity Functional Scale (LEFS)** | Orthopaedic | Lower Extremity Functional Scale | 36713869 | Measurement |
|  |  | Lower Extremity Functional Scale score | 36713866 | Observation |
| **Lysholm Scale** | Orthopaedic | Lysholm Knee Scoring Scale | 36713751 | Measurement |
|  |  | Lysholm Knee Scoring Scale score | 36713749 | Observation |
|  |  | Lysholm Knee Scoring Scale [LKSS] | 1988299 |  |
| **Marx Activity Scale** | Orthopaedic | Unmapped | | |
| **Maximal Inspiratory/Expiratory Pressure** | Neurologic | Peak expiratory pressure | 4066034 | Measurement |
|  |  | Peak inspiratory pressure | 4101694 |  |
|  |  | Inspiratory/expiratory ratio | 4119048 | Observation |
| **Mayo Elbow Performance Index (MEPI)** | Orthopaedic | Unmapped | | |
| **MDS-UPDRS- Revision** | Neurologic | Unified Parkinsons disease rating scale score | 44792308 | Measurement |
|  |  | Unified Parkinson's Disease Rating Scale (UPDRS) panel | 46236407 |  |
| **Mini Balance Evaluation Systems Test (Mini BESTest)** | Neurologic | Unmapped | | |
| **Modified Ashworth Scale** | Neurologic | Modified Ashworth Scale | 35623426 | Measurement |
|  |  | Modified Ashworth Scale score | 36685501 | Observation |
| **Modified Fatigue Impact Scale** | Neurologic | Fatigue impact scale | 40480572 | Measurement |
|  |  | Fatigue impact scale score | 40489875 | Observation |
| **Montreal Cognitive Assessment** | Neurologic | Montreal cognitive assessment | 44808666 | Measurement |
|  |  | Montreal Cognitive Assessment version 7.2 | 606672 |  |
|  |  | Montreal Cognitive Assessment version 7.3 | 606670 |  |
|  |  | Montreal Cognitive Assessment version 7.1 | 606673 |  |
|  |  | Montreal Cognitive Assessment version 8.1 | 606671 |  |
|  |  | Montreal cognitive assessment score | 36684973 | Observation |
|  |  | Montreal Cognitive Assessment [MoCA] | 43054876 |  |
| **Morton Mobility Index** | Orthopaedic | Unmapped | | |
| **Moss Attention Rating Scale** | Neurologic | MARS - Moss Attention Rating Scale | 37399409 | Measurement |
|  |  | Moss Attention Rating Scale score | 37396659 | Observation |
| **Multiple Sclerosis Functional Composite** | Neurologic | Unmapped | | |
| **Multiple Sclerosis Impact Scale** | Neurologic | Unmapped | | |
| **Multiple Sclerosis International Quality of Life Questionnaire** | Neurologic | Unmapped | | |
| **Multiple Sclerosis Quality of Life** | Neurologic | Unmapped | | |
| **Multiple Sclerosis Quality of Life Inventory** | Neurologic | Unmapped | | |
| **Neck Disability Index (NDI)** | Orthopaedic | Neck disability index | 44802398 | Measurement |
|  |  | Neck disability index score | 40487389 | Observation |
|  |  | Neck Disability Index [NDI] | 21493484 |  |
| **New Mobility Score** | Orthopaedic | Unmapped | | |
| **Numeric Pain-Rating Scale (NPRS) ^b^** | Orthopaedic | Pain intensity rating scale | 4137083 | Measurement |
|  |  | Visual analog pain scale | 4165600 |  |
|  |  | Visual analog scale pain score | 40481449 | Observation |
| **Örebro Musculoskeletal Pain Questionnaire** | Orthopaedic | Unmapped | | |
| **Oswestry Disability Index (ODI)** | Orthopaedic | Oswestry disability index | 4165295 | Measurement |
|  |  | Oswestry disability index score ODI | 1616831 |  |
|  |  | Oswestry Disability Index | 1617416 | Observation |
|  |  | Oswestry disability index score | 40488389 |  |
| **Pain Catastrophizing Scale** | Orthopaedic | Pain Catastrophizing Scale | 37397272 | Measurement |
|  |  | Pain Catastrophizing Scale score | 37399005 | Observation |
| **Pain Self-Efficacy** | Orthopaedic | Pain Self-efficacy Questionnaire | 37017917 | Measurement |
|  |  | Pain self efficacy questionnaire score | 40488834 | Observation |
| **Pain Visual Analogue Scale (VAS)** | Orthopaedic | Pain | 4329041 | Condition |
|  |  | Pain | 45881352 | Meas Value |
|  |  | Pain severity [Score] Visual analog score | 3036453 | Measurement |
|  |  | Visual analog pain scale | 4165600 |  |
|  |  | Visual analog scale pain score | 40481449 | Observation |
| **Parkinson's Fatigue Scale** | Neurologic | Unmapped | | |
| **Participation Assessment with Recombined Tools- Objective** | Neurologic | Unmapped | | |
| **Patient Specific Functional Scale (PSFS)** | Orthopaedic | Patient-Specific Functional Scale | 36714047 | Measurement |
|  |  | Patient-Specific Functional Scale score | 36714045 | Observation |
| **Patient-Rated Tennis Elbow Evaluation (PRTEE)** | Orthopaedic | Unmapped | | |
| **PDQ-39** | Neurologic | Unmapped | | |
| **PDQ-8** | Neurologic | Unmapped | | |
| **Pelvic Girdle Questionnaire (PGG)** | Orthopaedic | Unmapped | | |
| **Pierrynowski Questionnaire (EPQ)** | Orthopaedic | Unmapped | | |
| **Postural Assessment Scale for Stroke Patients** | Neurologic | PASS - Postural Assessment Scale for Stroke Patients | 42537354 | Measurement |
|  |  | PASS (Postural Assessment Scale for Stroke Patients) score | 42537357 | Observation |
| **PRO Function** | Orthopaedic | Unmapped | | |
| **PROMIS Anxiety** | Orthopaedic | PROMIS-29 Anxiety score | 42869723 | Observation |
|  |  | PROMIS emotional distress - anxiety - version 1.0 Tscore | 46236766 |  |
|  |  | PROMIS item bank - emotional distress - anxiety - version 1.0 | 40764683 |  |
|  |  | PROMIS short form - emotional distress - anxiety 4a - version 1.0 | 46235827 |  |
|  |  | PROMIS short form - emotional distress - anxiety 7a - version 1.0 | 40764951 |  |
|  |  | PROMIS short form - emotional distress - anxiety 4a - version 1.0 raw score | 46236681 |  |
|  |  | PROMIS short form - emotional distress - anxiety 7a - version 1.0 raw score | 46236696 |  |
|  |  | PROMIS-29 Anxiety score T-score | 42869722 |  |
| **PROMIS Depression** | Orthopaedic | PROMIS-29 Depression score | 42869721 | Observation |
|  |  | PROMIS-29 Depression score T-score | 42869720 |  |
|  |  | PROMIS emotional distress - depression - version 1.0 Tscore | 46236765 |  |
|  |  | PROMIS item bank - emotional distress - depression - version 1.0 | 40764713 |  |
|  |  | PROMIS short form - emotional distress - depression 4a - version 1.0 | 46235826 |  |
|  |  | PROMIS short form - emotional distress - depression 8b - version 1.0 | 40764952 |  |
|  |  | PROMIS short form - emotional distress - depression 4a - version 1.0 raw score | 46236682 |  |
| **PROMIS Fatigue** | Orthopaedic | PROMIS fatigue - version 1.0 Tscore | 46236768 | Observation |
|  |  | PROMIS-29 Fatigue score | 42869719 |  |
|  |  | PROMIS item bank - fatigue - version 1.0 | 40764556 |  |
|  |  | PROMIS-29 Fatigue score T-score | 42869718 |  |
|  |  | PROMIS short form - fatigue 4a - version 1.0 | 46235825 |  |
|  |  | PROMIS short form - fatigue 7a - version 1.0 | 40764953 |  |
|  |  | PROMIS short form - fatigue 4a - version 1.0 raw score | 46236683 |  |
|  |  | PROMIS short form - fatigue 7a - version 1.0 raw score | 46236694 |  |
| **PROMIS Pain Interference** | Orthopaedic | PROMIS pain interference - version 1.1 Tscore | 1176414 | Observation |
|  |  | PROMIS-29 Pain interference score | 42869717 |  |
|  |  | PROMIS pain interference - version 1.0 Tscore | 46236769 |  |
|  |  | PROMIS item bank - pain interference - version 1.1 | 1176315 |  |
|  |  | PROMIS item bank - pain interference - version 1.0 | 40764514 |  |
|  |  | PROMIS cancer pain interference - version 1.1 T-score | 1175731 |  |
|  |  | PROMIS-29 Pain interference score T-score | 42869716 |  |
|  |  | PROMIS short form - pain interference 6a - version 1.0 | 1175549 |  |
|  |  | PROMIS short form - pain interference 4a - version 1.0 | 1176430 |  |
|  |  | PROMIS short form - pain interference 6b - version 1.0 | 40764954 |  |
|  |  | PROMIS short form - pain interference 8a - version 1.0 | 46235683 |  |
|  |  | PROMIS short form - pain interference 6a - version 1.0 raw score | 1175856 |  |
|  |  | PROMIS short form - pain interference 4a - version 1.0 raw score | 1175435 |  |
|  |  | PROMIS short form - pain interference 6b - version 1.0 raw score | 46236693 |  |
|  |  | PROMIS short form - pain interference 8a - version 1.0 raw score | 46236684 |  |
| **PROMIS Physical Function** | Orthopaedic | PROMIS physical function - version 1.2 Tscore | 46236610 | Observation |
|  |  | PROMIS physical function - version 1.0 Tscore | 46236770 |  |
|  |  | PROMIS physical function - version 2.0 T-score | 37020016 |  |
|  |  | PROMIS item bank - physical function - version 1.0 | 40764348 |  |
|  |  | PROMIS item bank - physical function - version 2.0 | 37020102 |  |
|  |  | PROMIS-29 Physical function score T-score | 42869714 |  |
|  |  | PROMIS-29 Physical function score | 42869715 |  |
|  |  | PROMIS short form - physical function 10a - version 1.0 | 40764960 |  |
|  |  | PROMIS short form - physical function 10a - version 2.0 | 37021295 |  |
|  |  | PROMIS short form - physical function 6b - version 1.2 | 46235421 |  |
|  |  | PROMIS short form - physical function 4a - version 1.0 | 46235224 |  |
|  |  | PROMIS short form - physical function 20a - version 1.0 | 46235226 |  |
|  |  | PROMIS short form - physical function 10b - version 2.0 | 37021223 |  |
|  |  | PROMIS short form - physical function 8b - version 1.2 | 46235422 |  |
|  |  | PROMIS cancer item bank - physical function - version 1.1 | 1175443 |  |
|  |  | PROMIS short form - physical function 10b - version 2.0 raw score | 37020094 |  |
|  |  | PROMIS short form - physical function 8b - version 1.2 raw score | 46236591 |  |
|  |  | PROMIS short form - physical function w mobility aids - version 1.0 raw score | 21492093 |  |
| **PROMIS Physical Function UE** | Orthopaedic | PROMIS short form - upper extremity 7a - version 2.0 raw score | 37021089 | Observation |
|  |  | PROMIS short form - upper extremity 7a - version 2.0 | 37020608 |  |
|  |  | PROMIS upper extremity version 2.0 T-score | 37019789 |  |
| **PROMIS Social Roles** | Orthopaedic | PROMIS-29 Satisfaction with participation in social roles score | 42869713 | Observation |
|  |  | PROMIS satisfaction with social roles and activities - version 2.0 Tscore | 46236715 |  |
|  |  | PROMIS-29 Satisfaction with participation in social roles score T-score | 42869712 |  |
|  |  | PROMIS item bank - satisfaction with social roles and activities - version 2.0 | 46235397 |  |
|  |  | PROMIS short form - satisfaction with social roles and activities 6a - version 2.0 | 46235416 |  |
|  |  | PROMIS short form - satisfaction with social roles and activities 4a - version 2.0 | 46235415 |  |
|  |  | PROMIS short form - satisfaction with participation in social roles 4a - version 1.0 | 46235278 |  |
|  |  | PROMIS ability to participate in social roles and activities - version 2.0 Tscore | 46236714 |  |
|  |  | PROMIS short form - satisfaction with participation in social roles 7a - version 1.0 | 40764957 |  |
|  |  | PROMIS item bank - ability to participate in social roles and activities - version 2.0 | 46235410 |  |
|  |  | PROMIS short form - satisfaction with social roles and activities 8a - version 2.0 | 46235417 |  |
|  |  | PROMIS short form - satisfaction with participation in social roles 8a - version 1.0 | 46235225 |  |
|  |  | PROMIS short form - ability to participate in social roles and activities 8a - version 2.0 | 46235299 |  |
|  |  | PROMIS short form - satisfaction with social roles and activities 6a - version 2.0 raw score | 46236597 |  |
|  |  | PROMIS short form - satisfaction with social roles and activities 4a - version 2.0 raw score | 46236598 |  |
|  |  | PROMIS short form - satisfaction with participation in social roles 4a - version 1.0 raw score | 46236676 |  |
|  |  | PROMIS short form - satisfaction with participation in social roles 7a - version 1.0 raw score | 46236690 |  |
|  |  | PROMIS short form - satisfaction with social roles and activities 8a - version 2.0 raw score | 46236596 |  |
|  |  | PROMIS short form - satisfaction with participation in social roles 8a - version 1.0 raw score | 46236679 |  |
|  |  | PROMIS short form - ability to participate in social roles and activities 8a - version 2.0 raw score | 46236605 |  |
| **Purdue Pegboard** | Orthopaedic | Purdue pegboard scale | 4158641 | Measurement |
| **Quebec Back Pain Disability Scale** | Orthopaedic | Unmapped | | |
| **Rancho Levels of Cognitive Function** | Neurologic | Rancho Los Amigos Levels of Cognitive Functioning Scale | 36714643 | Measurement |
|  |  | Rancho Los Amigos Levels of Cognitive Functioning Scale score | 37396840 | Observation |
| **Readiness for Return-to-Work Scale** | Orthopaedic | Unmapped | | |
| **Reintegration to Normal Living** | Neurologic | Unmapped | | |
| **Rivermead Mobility Index** | Neurologic | Rivermead Mobility Index | 44811906 | Measurement |
|  |  | Rivermead Mobility Index score | 44814117 | Observation |
| **Roland-Morris Disability Questionnaire** | Orthopaedic | Roland-Morris disability questionnaire score | 44789320 | Measurement |
|  |  | RMDQ (Roland-Morris Disability Questionnaire) score | 36685580 | Observation |
|  |  | Roland Morris Disability Questionnaire panel [RMDQ] | 1989100 |  |
| **Roles and Maudsley Score** | Orthopaedic | Unmapped | | |
| **Roll Test** | Neurologic | Unmapped | | |
| **Romberg** | Neurologic | Romberg test | 4162447 | Procedure |
| **Satisfaction with Life Scale** | Neurologic | Satisfaction with life scale | 4121656 | Measurement |
| **Self-Paced Walk** | Orthopaedic | Walk in room - self-performance during assessment period [CMS Assessment] | 3045663 | Observation |
|  |  | Walk in corridor - self-performance during assessment period [CMS Assessment] | 3047307 |  |
| **Sensory Organization Test** | Neurologic | Sensory organization test | 36684868 | Measurement |
| **Sharpened Romberg** | Neurologic | Unmapped | | |
| **Short Form Health Survey of the Medical Outcome Study ^b^** | Neurologic | Medical outcomes study short form general health survey | 4169170 | Measurement |
|  |  | Medical outcomes study short form general health survey - 20 | 4157407 |  |
|  |  | Medical outcomes study short form general health survey - 36 | 4165282 |  |
|  |  | MOS SF-20 (Medical Outcomes Study 20-Item Short Form Health Survey) score | 35610202 | Observation |
|  |  | MOS SF-36 (Medical Outcomes Study 36-Item Short Form Health Survey) score | 35610204 |  |
| **Short Physical Performance Battery** | Orthopaedic | Short Physical Performance Battery | 37017915 | Measurement |
|  |  | Short Physical Performance Battery score | 37017960 | Observation |
| **Shoulder Pain And Disability Index (SPADI)** | Orthopaedic | Unmapped | | |
| **Sidelying Test** | Neurologic | Unmapped | | |
| **Single-Leg Hop Tests** | Orthopaedic | Unmapped | | |
| **Star Excursion Balance Test** | Orthopaedic | Unmapped | | |
| **Step Test** | Orthopaedic | Unmapped | | |
| **Stroke Impact Scale** | Neurologic | Stroke impact scale version 3.0 | 45767552 | Measurement |
|  |  | Stroke impact scale version 3.0 score | 45772944 | Observation |
| **Tampa Scale of Kinesiophobia** | Orthopaedic | Unmapped | | |
| **Tardieu Spasticity Scale (Modified Tardieu)** | Neurologic | Unmapped | | |
| **Tegner Activity Scale** | Orthopaedic | Tegner Activity Scale | 3654720 | Measurement |
|  |  | Tegner Activity Scale panel [Tegner] | 1988348 | Observation |
| **The America Shoulder and Elbow Surgeons Shoulder Scale (ASES)** | Orthopaedic | Unmapped | | |
| **The Copenhagen Hip and Groin Outcome Score (HAGOS)** | Orthopaedic | Unmapped | | |
| **The Hip Disability and Osteoarthritis Outcome Score (HOOS)** | Orthopaedic | Hip disability and osteoarthritis outcome score | 40493275 | Measurement |
|  |  | Hip disability and osteoarthritis outcome score | 40486510 | Observation |
|  |  | Hip Dysfunction and Osteoarthritis Outcome Score [HOOS] | 42869847 |  |
|  |  | Hip disability and osteoarthritis outcome score - physical function shortform panel [HOOS-PS] | 1988793 |  |
| **The Hip Outcome Score (Hos)** | Orthopaedic | Unmapped | | |
| **The International Hip Outcome Tool (IHOT-33)** | Orthopaedic | Unmapped | | |
| **The Modified Harris Hip Score (MHHS)** | Orthopaedic | Modified Harris hip score | 40486424 | Measurement |
|  |  | Modified Harris hip score | 40486420 | Observation |
| **The Subgroups for Targeted Treatment Back Screening Tool (STarT Back)** | Orthopaedic | Subgroups for targeted treatment back screening tool | 40491838 | Measurement |
|  |  | Subgroups for targeted treatment back screening tool score | 40491837 |  |
|  |  | STarT (Subgroups for Targeted Treatment) Back Screening Tool subscore | 37310735 |  |
|  |  | STarT Back Screening Tool panel | 37021490 | Observation |
|  |  | STarT Back Screening risk level | 37020073 |  |
| **Timed 25 Foot Walk** | Neurologic | Unmapped | | |
| **Timed Single-Leg Stance** | Orthopaedic | One leg stand test time | 45768778 | Observation |
| **Timed Up and Go (TUG)** | Neurologic | Timed up and go mobility test | 40480727 | Measurement |
|  |  | Timed up and go mobility test score | 40480298 | Observation |
| **Timed Up and Go (Tug) Dual Task** | Neurologic | Unmapped | | |
| **Trunk Impairment Scale** | Neurologic | Unmapped | | |
| **Trunk Muscle Power and Endurance** | Orthopaedic | Unmapped | | |
| **Unipedal Stance Test** | Neurologic | Unmapped | | |
| **Victorian Institute of Sport Assessment-Achilles** | Orthopaedic | Victorian Institute of Sport Assessment-Achilles questionnaire | 40486426 | Measurement |
| **Victorian Institute of Sport Assessment-Achilles** | Orthopaedic | Victorian Institute of Sport Assessment-Achilles questionnaire score | 40486002 | Observation |
| **Visual Analog Scale (VAS)** | Orthopaedic | Visual analog scale | 4158877 | Measurement |
|  |  | Visual analog scale score | 40488832 | Observation |
| **Visual Analog Scale for Fatigue** | Neurologic | Unmapped | | |
| **Western Ontario and McMaster Universities Osteoarthritis Index (WOMAC)** | Orthopaedic | Western Ontario and McMaster Universities osteoarthritis index | 42873025 | Measurement |
| **WHO Quality of Life - Bref** | Neurologic | Unmapped | | |
| **Wolf Motor Function Test** | Neurologic | Unmapped | | |
| **Work and Health Questionnaire** | Orthopaedic | Unmapped | | |
| ^a^ Although these measures can be used in many populations, the category indicated here is based on which Evaluation Database to Guide Effectiveness (EDGE) document(s) or clinical practice guideline(s) the assessment was listed on.  ^b^ These assessments were initially identified on both the neurologic and orthopaedic assessment list; however, there were only included in the analysis for the category listed in this table. | | | | |
